# How and why do Australians obtain blood pressure devices for use at home? A mixed-methods study

**DOI:** 10.1101/2025.02.27.24318446

**Authors:** Eleanor Clapham, Samuel Carmichael, Dean S Picone, Aletta E Schutte, Kaylee Slater, John Stevens, Mark R Nelson, Markus Schlaich, Rachel E Climie, Ross T. Tsuyuki, George Stergiou, Norm RC Campbell, Niamh Chapman

## Abstract

**Background:** Only 10-20% of blood pressure (BP) devices available are validated. Little is known about how and why patients choose BP devices for home BP measurement (HBPM), which was the aim of this study.

**Methods:** Mixed-methods study (online survey (n=241), phone interviews among a purposive subsample (n=27)) among adults who perform HBPM in Australia (June-Dec 2023). Survey questions determined how BP devices were obtained, device make/model and factors influencing device selection. Interviews further explored these topics. Device validation status was determined using the STRIDE BP and Medaval websites.

**Results:** Participants were middle aged (58±16 years, 52% women) and 91% purchased a device for HBPM (n=189; 9% borrowed a device), with 69% (n=130) purchased from pharmacies (53% validated) and 21% (n=39) purchased online (51% validated).

Accuracy was said to be the most important consideration when choosing a device for most participants (n=129, 77%). Interview participants described using brand recognition, online reviews and cost to select an ‘accurate’ device; avoiding cheaper devices and preferring brands used in healthcare settings. Participants did not consider validation status and did not receive advice on device accuracy at point-of-sale.

**Conclusion:** This study highlights real world experiences of adults when obtaining HBPM devices that can be used to inform strategies to direct adults to validated devices. Strategies such as increasing signage at the point-of-sale and training healthcare practitioners to identify and direct consumers to validated devices may be effective in increasing uptake. Regulatory bodies should mandate the sale of validated devices in healthcare settings to increase availability.

**GRAPHICAL ABSTRACT:** 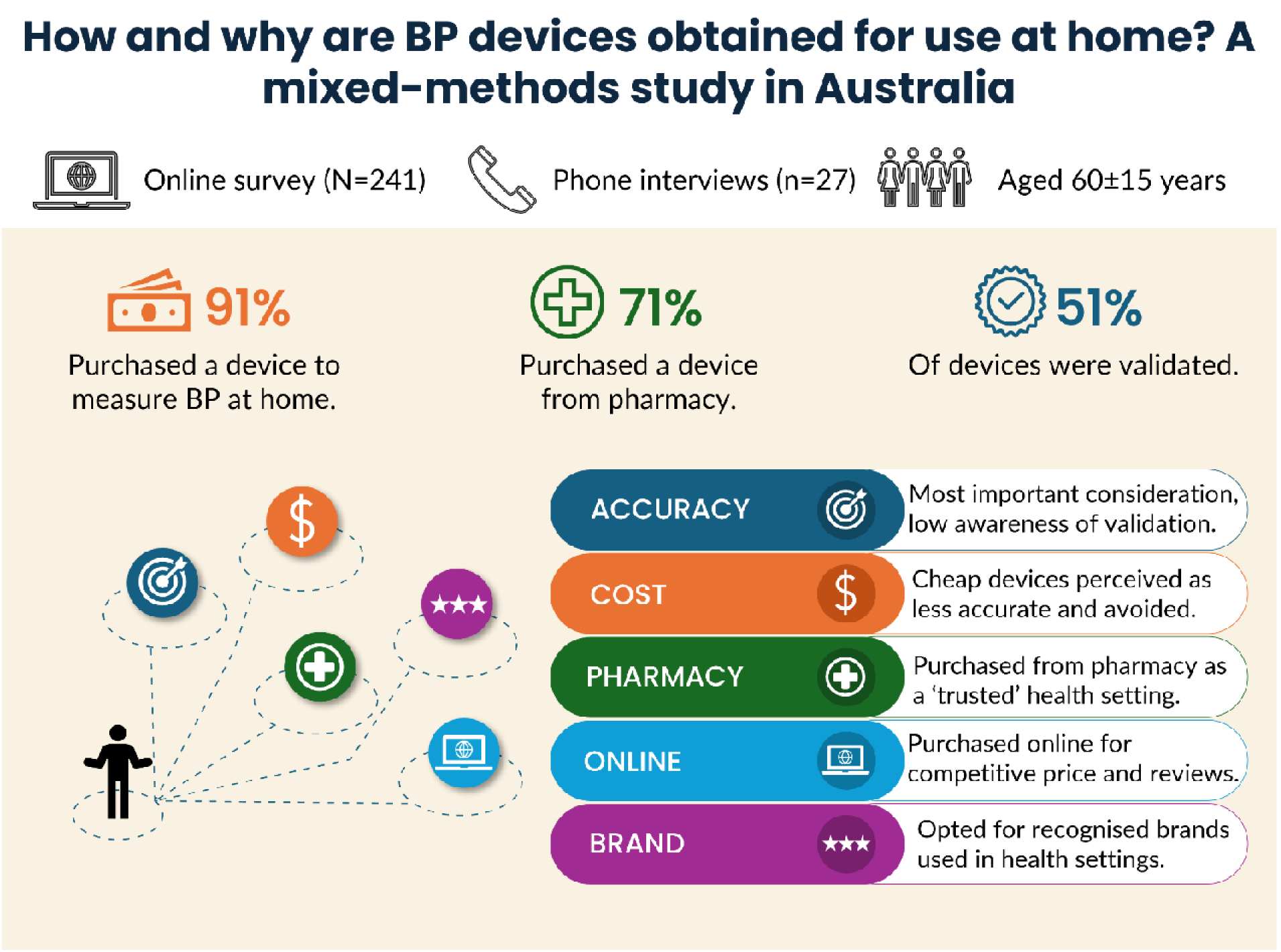

## INTRODUCTION

High blood pressure (BP) or hypertension is the leading modifiable risk factor for morbidity and mortality worldwide (1, 2). Traditionally, high BP is detected through in-clinic BP measurement by a healthcare practitioner, but there are many well established limitations of this method (3, 4). As such, home BP measurement (self-monitored BP; HBPM) using a clinically validated, automated, upper-arm cuff-based device is recommended by international clinical guidelines as one method for hypertension diagnosis and long-term BP monitoring (5–7). The key first step to enable adults to perform HBPM is obtaining a validated BP device (4, 7).

Clinically validated BP devices are those which have evidence of passing accuracy testing according to an internationally accepted protocol (8). Clinically validated BP devices provide reliably accurate BP readings to appropriately inform BP management (9–12). However, evidence of passing clinical validation is rarely required by regulatory bodies for the sale of BP devices, with such regulations focussing on the safety of medical devices (13–15).

At the global level, previous research identified that <25% of home BP devices available for purchase are validated, and validated devices are more expensive than devices without evidence of validation (16, 17), which may be a barrier to use of validated BP devices among consumers. However, studies of adults who own BP devices for HBPM report that between 20-81% of devices used by participants were validated (11, 18–21). These data indicate that despite the low availability and higher cost of validated BP devices there is wide variation in the use of validated BP devices. Little is known about the strategies used by consumers to select a BP device, which could inform future strategies to support consumers to obtain a validated device for use at home.

Recent efforts have focused on raising awareness of the low market availability of validated BP devices. However, there is a critical knowledge gap in the experience of consumers when obtaining a device for HBPM and the factors that influence device selection. As such, this study aimed to determine if adults who measure BP at home use a validated BP device for HBPM, how their BP device used for HBPM was obtained and understand the factors that influence BP device selection.

## METHODS

### Study design

A mixed methods study with a 30-item online survey and optional semi-structured phone interviews among Australian adults who measure BP at home was conducted between June and December 2023. The study was undertaken to determine the quality of HBPM and barriers or enablers to high-quality HBPM. The full methods of the study have been published in detail previously (22). For this present paper, the online survey provided basic demographic and health information, the BP device used (make and model), how and where the BP device was obtained and factors influencing device selection (Supplementary Table S1). Interviews explored the experience of participants when obtaining a BP device, the factors considered and barriers and enablers to obtaining a validated device (Supplementary Table S2). All participants completed the survey electronically via the secure online platform ‘Research Electronic Data Capture’ (REDCap) and interviews were conducted using Microsoft Teams (23, 24). This study was approved by the Human Research Ethics Committee of the University of Tasmania (H0028867) and informed consent was provided by all participants prior to participation.

### Participant recruitment

The survey was conducted nationally within Australia and advertised by supporting organisations (e.g., Hypertension Australia, Stroke Foundation), and healthcare organizations and the research team via email, social media advertisements, newsletters, physical flyers, and digital notice boards. Participants had the opportunity to enter a prize draw to win one of 30 $100 e-gift vouchers as compensation for their time. Those who consented to follow-up interviews were recruited using an information power purposive sampling approach (25), to ensure a diverse range of participant characteristics based on location, gender, education level, health literacy, and BP device validation status. Participants were recruited until data saturation had been reached (26), which was decided based on regular review of emerging themes within interview data in consultation with an experienced qualitative researcher (NC).

### Inclusion and exclusion criteria

Adults (≥18 years), who lived in Australia, and had measured BP at home using an upper-arm or wrist cuff BP device at least once within in the previous 12 months were eligible to take part. Participants who did not provide BP device brand and/or model number were excluded from the current analysis (Figure 1).

**Figure 1.**
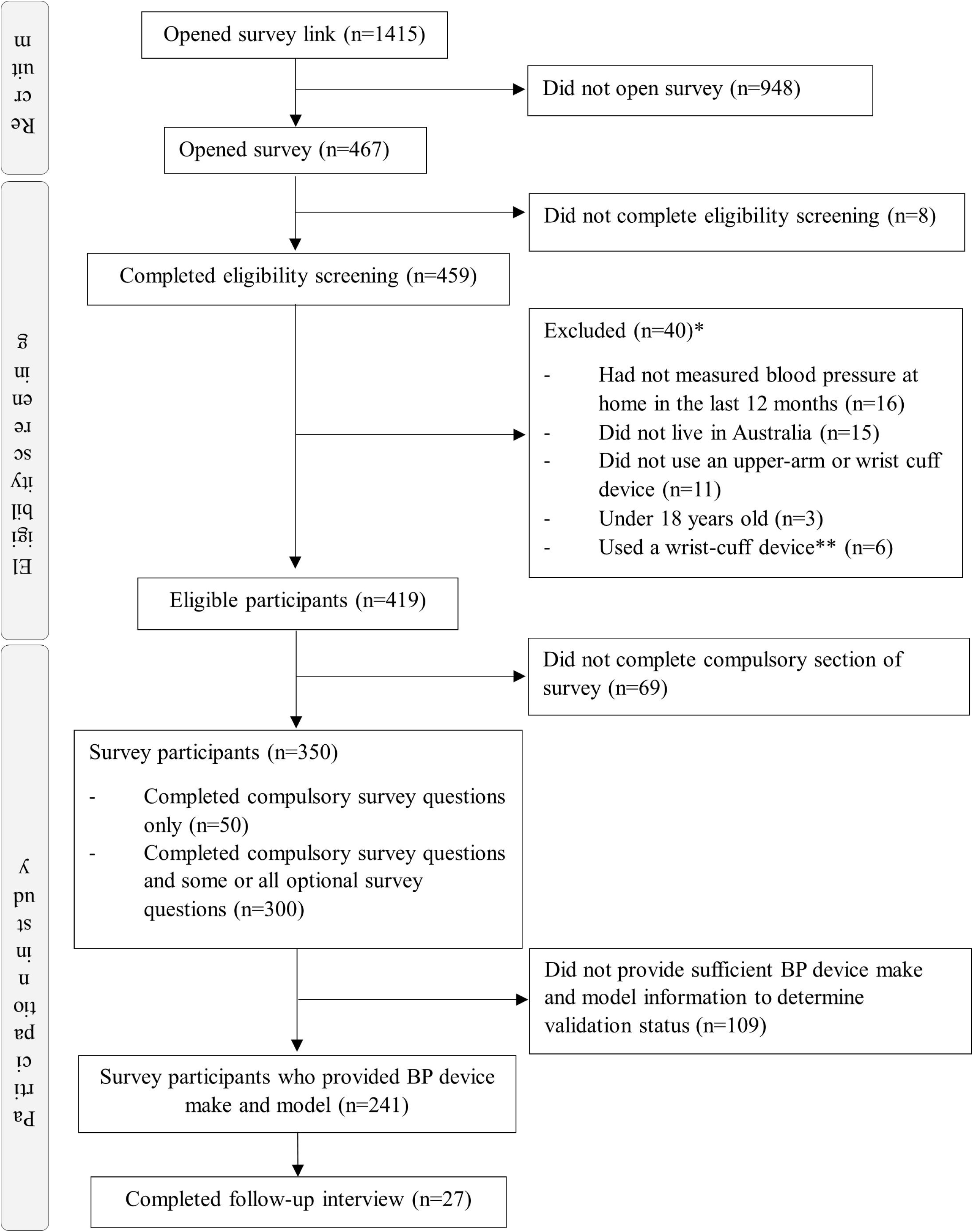
Participant flow diagram. *Sum of individual exclusion criteria equals >40 as some excluded participants met more than one exclusion criteria. **The use of a wrist cuff device was removed as an exclusion criterion after the release of the survey.

### Demographic information

Demographic data from participants included age, gender, ethnicity, location, and education. Postcode was used to determine geographical remoteness using the Modified Monash Model (27) and socioeconomic status via the Index of Relative Socio-Economic Advantage and Disadvantage (ISRAD) (28).

### Obtaining a device for home BP measurement

This survey covered 11 mandatory questions relating to acquiring a home BP device, including how and where the BP device was obtained, factors considered when selecting a BP device and device make/model (Supplementary Table S1).

Semi-structured interviews were conducted to further explore experiences and perspectives of participants when selecting a BP device, support and/or information received to obtain a BP device, and barriers and enablers experienced during this process (Supplementary Table S2). Interviews were conducted online, recorded and auto-transcribed verbatim.

### Device validation status

Device validation status was determined using STRIDE BP (29) and Medaval (30) websites using the method outlined by Picone et. al. (31). The BP device make and model number provided by survey participants was input into each website and validation status recorded. BP devices classified as ‘validated’ and ‘equivalent’ were categorised as validated, and BP devices classified as ‘non-validated’ or not present on either website were classified as ‘non-validated’.

### Device cost

To obtain BP device cost, the cost of all BP devices as advertised on the online pharmacies of Terry White Chemist, Blooms the Chemist, Chemist Warehouse, Better Value Pharmacy, and Amcal was recorded (EC) (01/12/2024). The lowest recorded cost across online pharmacies for each BP device was used for analysis. Data were extracted in AUD and converted to USD using the exchange rate on 01/12/2024 (0.66850). This methodology was informed by the purchasing strategy described by interview participants and is consistent with an approach published previously (16). Cost data of participants who borrowed the BP device used for HBPM were excluded from analysis.

### Factors contributing to choosing a BP device

Survey participants were provided with six factors that may influence BP device selection: cost, quality, accuracy, additional features, recommendations from friends and family, and recommendations from medical professionals. Participants ranked how important each factor was to them when selecting a BP device on a 6-point Likert Scale, where 1=least important factor and 6=most important factor. For data analysis and presentation, the LIKERT scale options (1–6) were categorised as most important (options 6 & 5), neutral (options 3 & 4) and least important (options 1 & 2).

### Data analysis

Descriptive statistics were used to analyse demographic characteristics, and characteristics of BP devices, including cost and factors contributing to participant BP device selection analysed on Stata/SE (V18.0). Continuous data are presented as mean (SD) and categorical data are presented as n (%). Nonparametric equality-of-medians tests were used to determine if there was an association between the cost of validated and non-validated devices. Pearson’s chi-squared test was used to determine differences in demographic characteristics between participants who provided enough information to ascertain device validation status and those who did not, and participants who used a validated BP device and those who used a non-validated device. A p-value <0.05 was considered statistically significant.

De-identified verbatim interview transcripts were analysed on Nvivo (V 1.7.1 (1534)). Interview transcripts were analysed by deductive thematic analysis. The thematic analysis coding framework was constructed by SC to categorise participant answers according to the interview questions and their components (Supplementary Table S3). Four researchers (EC, SC, NC and DP) read a subset of transcripts (n=5) to ensure suitability of the coding framework. Once the coding framework was agreed, two study investigators (EC, SC) independent thematically analysed interview transcripts with iterative feedback with an experienced qualitative researcher (NC). A fifth independent reviewer (KS) analysed a subset of interviews (n=10) to ensure reported themes reflected the data. To support researcher reflexivity, researchers involved in thematic analysis included those experienced in qualitative research methods (NC), BP measurement methods (DP, NC), patient needs in health services (NC, KS), information delivery and BP management (NC, EC). Thematic analysis themes and illustrative quotes are outlined in Supplementary Table S4. Illustrative, de-identified verbatim quotes are presented throughout results as empirical data consistent with recommendations for the reporting of qualitative research (32).

## RESULTS

### Participant characteristics

Characteristics of survey participants (n=241) are described in Table 1, participant flow is shown in Figure 1 and characteristics of interview participants (n=27) are reported in Supplementary Table S6. Participants were middle aged (mean age 60±15 years), 52% were women, 90% were white, 49% were from metropolitan areas and a third were from areas of social disadvantage. Most participants had a diagnosis of hypertension (n=177, 73%) and 30% (n=61) had a previous heart attack or stroke.

**Table 1.**
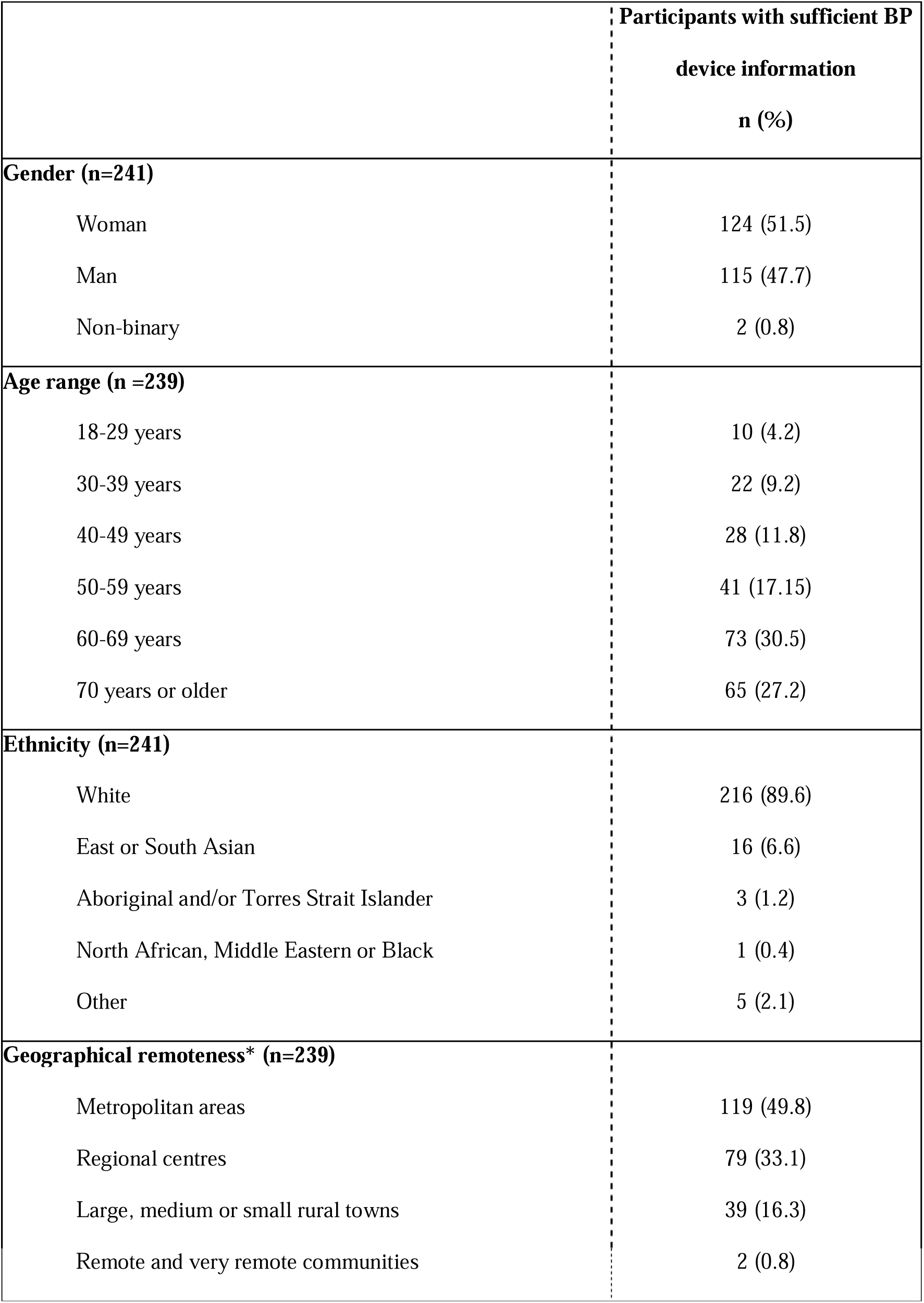
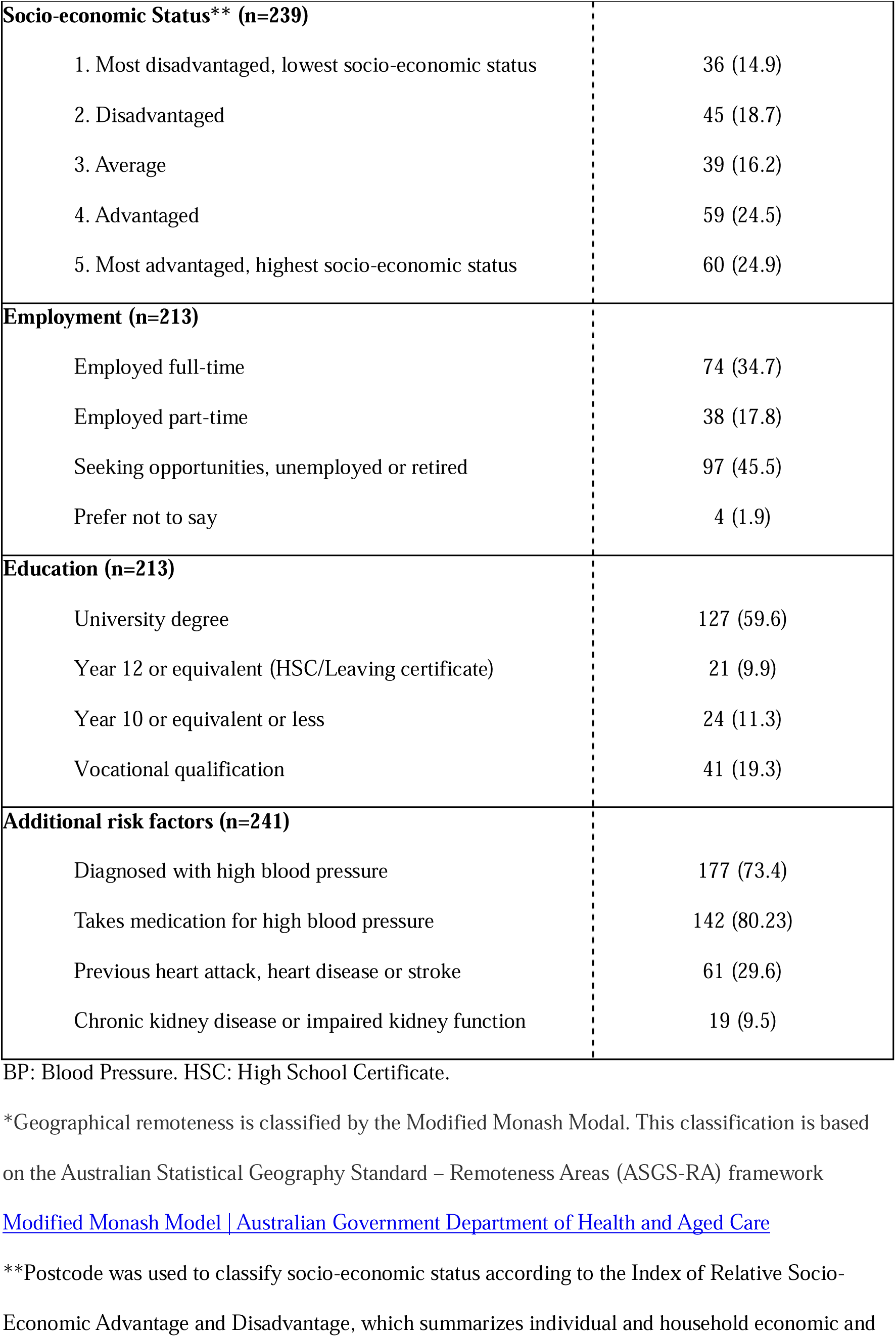
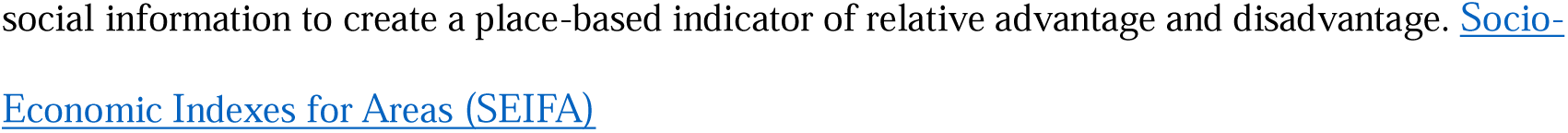
Characteristics of Australian adult study participants who measure BP at home (n=241).

|  | Participants with sufficient BP<br>device information<br>n (%) |
| --- | --- |
| **Gender (n=241)** |  |
| Woman | 124 (51.5) |
| Man | 115 (47.7) |
| Non-binary | 2 (0.8) |
| **Age range (n =239)** |  |
| 18-29 years | 10 (4.2) |
| 30-39 years | 22 (9.2) |
| 40-49 years | 28 (11.8) |
| 50-59 years | 41 (17.15) |
| 60-69 years | 73 (30.5) |
| 70 years or older | 65 (27.2) |
| **Ethnicity (n=241)** |  |
| White | 216 (89.6) |
| East or South Asian | 16 (6.6) |
| Aboriginal and/or Torres Strait Islander | 3 (1.2) |
| North African, Middle Eastern or Black | 1 (0.4) |
| Other | 5 (2.1) |
| **Geographical remoteness\* (n=239)** |  |
| Metropolitan areas | 119 (49.8) |
| Regional centres | 79 (33.1) |
| Large, medium or small rural towns | 39 (16.3) |
| Remote and very remote communities | 2 (0.8) |
| **Socio-economic Status\*\* (n=239)** |  |
| 1. Most disadvantaged, lowest socio-economic status | 36 (14.9) |
| 2. Disadvantaged | 45 (18.7) |
| 3. Average | 39 (16.2) |
| 4. Advantaged | 59 (24.5) |
| 5. Most advantaged, highest socio-economic status | 60 (24.9) |
| **Employment (n=213)** |  |
| Employed full-time | 74 (34.7) |
| Employed part-time | 38 (17.8) |
| Seeking opportunities, unemployed or retired | 97 (45.5) |
| Prefer not to say | 4 (1.9) |
| **Education (n=213)** |  |
| University degree | 127 (59.6) |
| Year 12 or equivalent (HSC/Leaving certificate) | 21 (9.9) |
| Year 10 or equivalent or less | 24 (11.3) |
| Vocational qualification | 41 (19.3) |
| **Additional risk factors (n=241)** |  |
| Diagnosed with high blood pressure | 177 (73.4) |
| Takes medication for high blood pressure | 142 (80.23) |
| Previous heart attack, heart disease or stroke | 61 (29.6) |
| Chronic kidney disease or impaired kidney function | 19 (9.5) |

BP: Blood Pressure. HSC: High School Certificate.
\*Geographical remoteness is classified by the Modified Monash Model. This classification is based on the Australian Statistical Geography Standard – Remoteness Areas (ASGS-RA) framework
Modified Monash Model | Australian Government Department of Health and Aged Care
\*\*Postcode was used to classify socio-economic status according to the Index of Relative Socio-Economic Advantage and Disadvantage, which summarizes individual and household economic and
social information to create a place-based indicator of relative advantage and disadvantage. Socio-Economic Indexes for Areas \(SEIFA\)

Participants that provided BP device data were older and had a higher prevalence of history of heart disease, heart attack or stroke (*p*<0.05) than those who did not provide BP device information (Supplementary Table S5).

### BP devices used by participants to measure BP at home

Most participants (n=232, 96%) used an upper-arm cuff device for HBPM (49% validated, Table 2). Only nine (4%) participants used a wrist-cuff device (67% validated). Some interviewees had considered using a wrist cuff BP device due to convenience but felt that upper-arm cuff devices were more accurate (Figure 2). *“I had heard that wrist ones, even though they looked like they’ll be a lot easier, aren’t as good as the upper arm one. So, I got an upper arm one”*.

**Figure 2.**
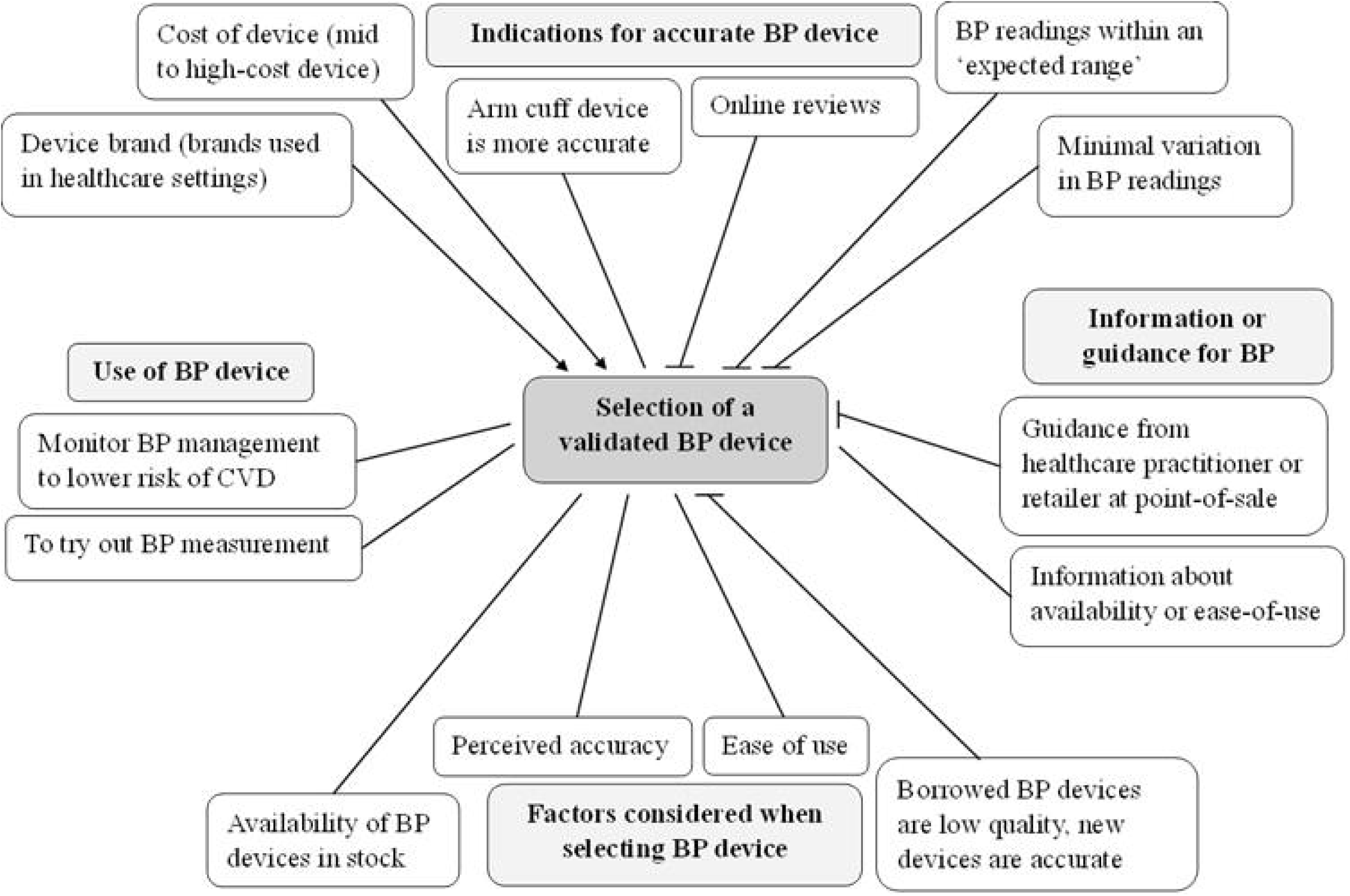
Factors impacting the selection of a validated BP device. Verbatim interview transcripts (n=27) were thematically analysed using a deductive thematic analysis coding framework. Arrow: Factor enabled participants to select a validated BP device. Blunt pointer: Factor did not enable participants to select a validated BP device. Pointer with no end: Factor was considered for BP device selection but was not relevant to purchase of a validated device. BP: Blood pressure, CVD: Cardiovascular disease

**Table 2.**
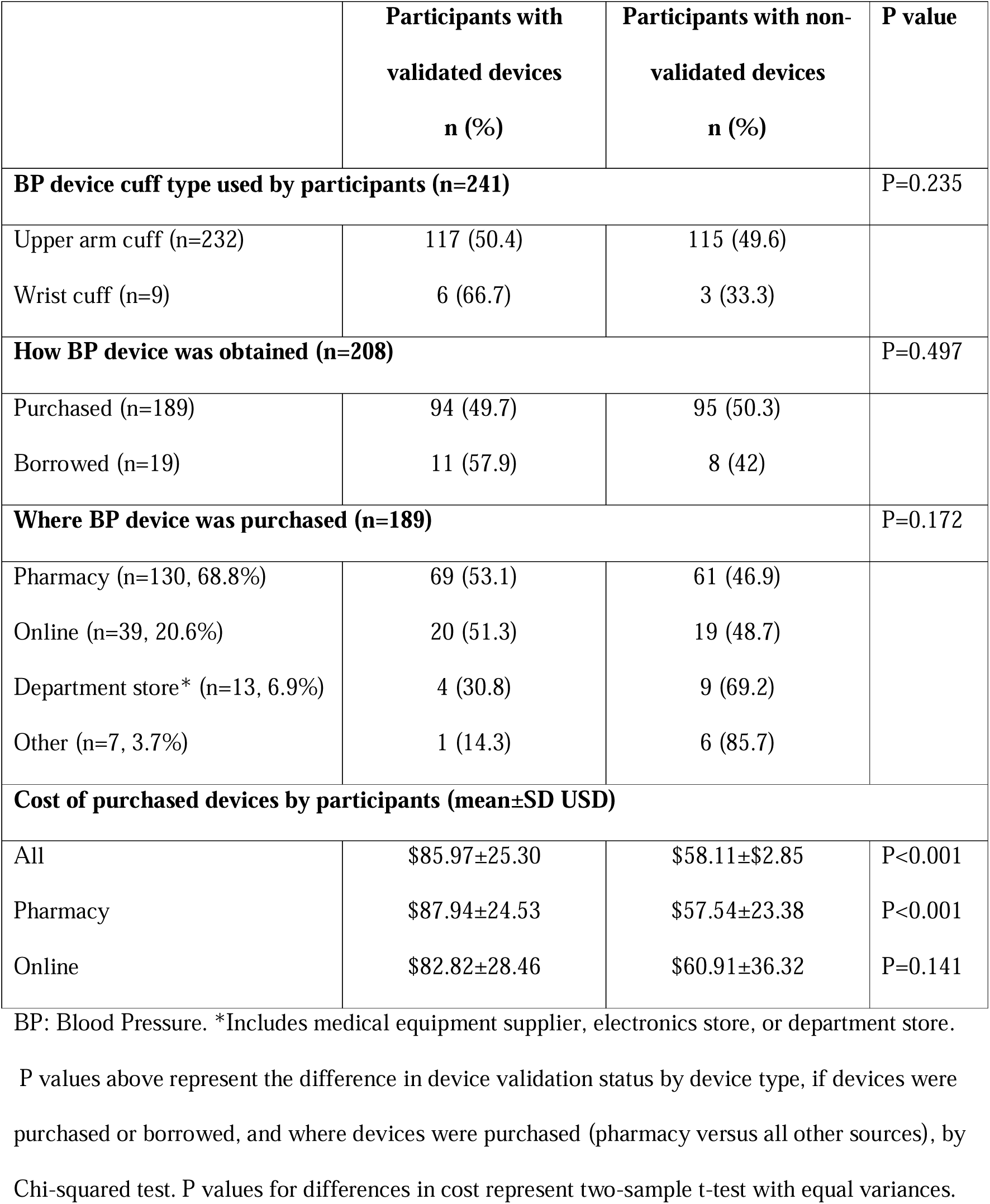
Characteristics and validation status of BP devices used by Australian adults who measure BP at home (n=241).

|  | **Participants with**<br>**validated devices**<br>**n (%)** | **Participants with non-**<br>**validated devices**<br>**n (%)** | **P value** |
| --- | --- | --- | --- |
| **BP device cuff type used by participants (n=241)** |  |  | P=0.235 |
| Upper arm cuff (n=232) | 117 (50.4) | 115 (49.6) |  |
| Wrist cuff (n=9) | 6 (66.7) | 3 (33.3) |  |
| **How BP device was obtained (n=208)** |  |  | P=0.497 |
| Purchased (n=189) | 94 (49.7) | 95 (50.3) |  |
| Borrowed (n=19) | 11 (57.9) | 8 (42) |  |
| **Where BP device was purchased (n=189)** |  |  | P=0.172 |
| Pharmacy (n=130, 68.8%) | 69 (53.1) | 61 (46.9) |  |
| Online (n=39, 20.6%) | 20 (51.3) | 19 (48.7) |  |
| Department store* (n=13, 6.9%) | 4 (30.8) | 9 (69.2) |  |
| Other (n=7, 3.7%) | 1 (14.3) | 6 (85.7) |  |
| **Cost of purchased devices by participants (mean±SD USD)** |  |  |  |
| All | \$85.97±25.30 | \$58.11±\$2.85 | P<0.001 |
| Pharmacy | \$87.94±24.53 | \$57.54±23.38 | P<0.001 |
| Online | \$82.82±28.46 | \$60.91±36.32 | P=0.141 |

BP: Blood Pressure. \*Includes medical equipment supplier, electronics store, or department store.
P values above represent the difference in device validation status by device type, if devices were purchased or borrowed, and where devices were purchased (pharmacy versus all other sources), by Chi-squared test. P values for differences in cost represent two-sample t-test with equal variances.

Among the 241 participants, 86 unique devices were used, the make and models of devices used by participants are outlined in Supplementary Table S7. The most used BP device among participants (n=59, 25%) was validated, while the second most used device (n=17, 7%) has no published evidence of validation.

### Obtaining a BP device

Most participants (91%) purchased a device for HBPM (50% validated) and few (n=19, 9%) had borrowed a device (58% validated, Table 2). Interview participants described that owning a device was important as this allowed for round-the-clock access to HBPM. Conversely, borrowing a BP device was described as an affordable way to try out HBPM, but inconvenient for long-term BP monitoring (Figure 2).

Among those that purchased a device, 69% purchased from pharmacies (53% validated) and 21% purchased online (51% validated). There was no difference in the proportion of participants who used a validated BP device according to the place the device was purchased, geographical remoteness, or relative socioeconomic status (Supplement Table S7).

### Factors considered when obtaining a BP device

Accuracy was said to be the most important consideration for most survey participants when choosing a BP device (77%, n=129, Figure 3), which was reiterated by interview participants (Figure 2). However, no interview participants were aware that clinical validation is a reliable indication of BP device accuracy, and many assumed all BP devices would be accurate. *“I just assumed that they were all accurate […] it’s new, it’s in a box, hasn’t been used, so it’ll give me an accurate result”*. Interviewees defined an accurate BP device as one that consistently produced minimal variation in BP readings, matched in-clinic BP readings, and had online reviews affirming reliability or accuracy (Figure 2).

**Figure 3.**
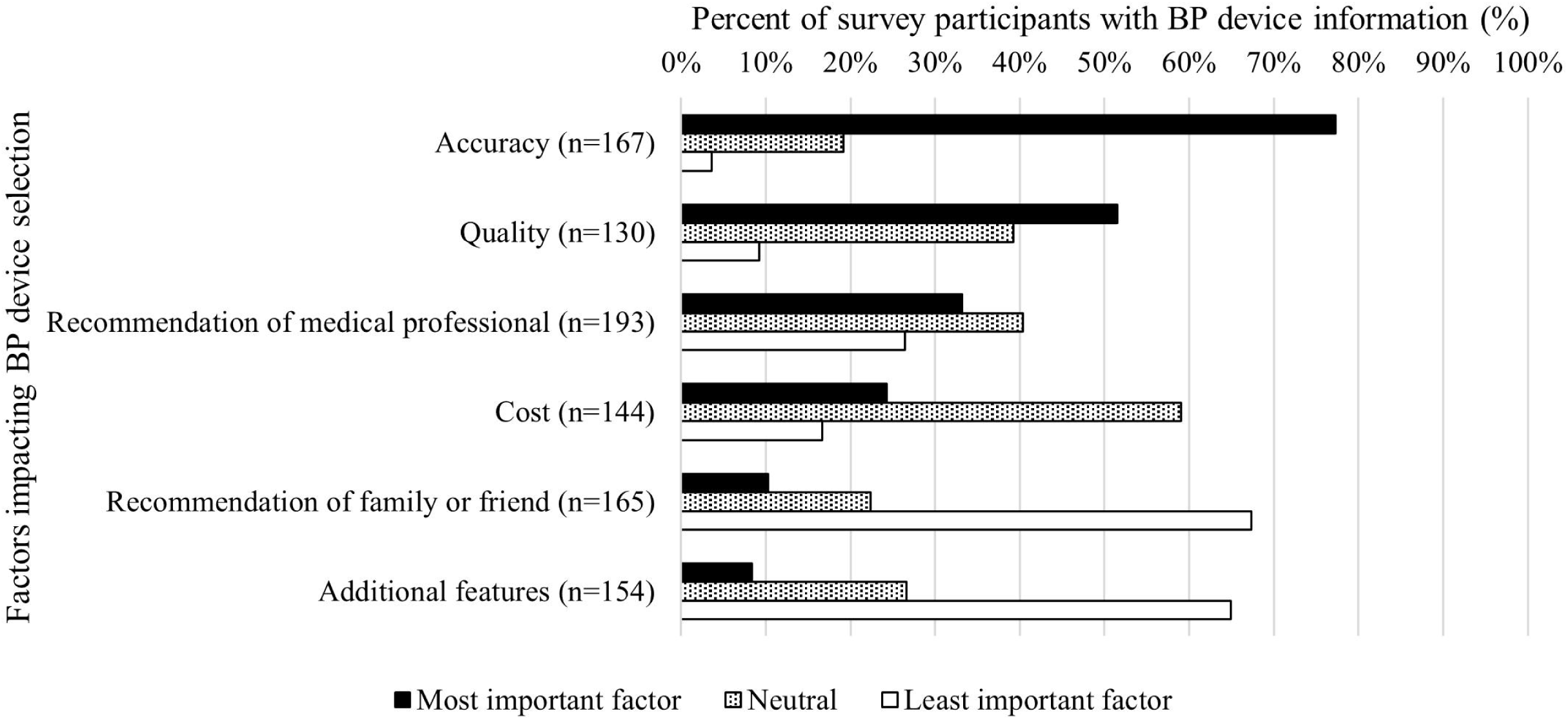
Factors contributing to participant BP device selection. Survey LIKERT scale options (1–6) have been categorised as most important (options 6 & 5), neutral (options 3 & 4) or least important (options 1 & 2). BP: Blood pressure.

Some interviewees described brand trust as an important factor when selecting a BP device. These participants actively sought out a particular brand of BP device that they had observed in healthcare settings and perceived it as trustworthy and accurate (Figure 2). *“It seems pretty similar brand to what the doctors are using, so I think it’s a reasonable product.”*.

A third of survey participants (n=64, 33%) considered recommendations from a medical professional to be the most important factor when selecting a BP device (Figure 3). However, interview participants described feeling unsupported in their decision-making when selecting a BP device. Interviewees rarely reported receiving specific recommendations for BP devices from their GP and explained that pharmacists present at point-of-sale focused on retail availability rather than device accuracy (Figure 2). *“They weren’t overly helpful or overly interested in helping, but then they may also have not been knowledgeable enough to help in any way. I mean you just you’re buying a product to them…and they’re just the retailer of it.”*.

### Cost of BP devices purchased by participants to measure BP at home

Cost (in AUD) of BP devices is reported in Supplementary Table S7, while cost in text is reported in USD. The mean cost of BP devices purchased by survey participants was $76.01±2.55 USD. The cost of validated devices ($85.97±25.30) was on average $27.86 USD more expensive than non-validated devices ($58.11±2.85, p=<0.001, Table 2). There was no significant difference in the cost of validated and unvalidated devices by devices purchased online, socioeconomic status or geographical location.

Nearly two-thirds of survey participants did not report cost as an important factor (59%, n=85, Figure 3) when obtaining a BP device. However, when discussed in more detail during interviews, interviewees described avoiding cheaper BP devices as they perceived them to be less accurate than more expensive options (Figure 2). Several interviewees recalled receiving financial reimbursement through their private healthcare fund, but felt this did not impact their choice of device but instead reduced the financial commitment of HBPM.

## DISCUSSION

In this study, we identified the factors that inform BP device selection among consumers who measure BP at home. Most participants purchased a BP device from a pharmacy and only half of the BP devices used by participants were validated. Accuracy was said to be the most important consideration for device selection however participants did not have any knowledge of validation status and did not receive guidance to select a validated BP device. Instead, participants selected brands used in healthcare settings and some avoided ‘cheap’ devices to select a BP device they perceived to be accurate. These findings can inform strategies to support consumers to select validated BP devices for use at home such as providing clear signage at point-of-sale and training healthcare practitioners about the need to use validated devices for BP measurement. These findings also highlight the role of government to support consumers through regulation on the sale of validated BP devices, particularly in health settings. A summary of recommendations to improve uptake of validated BP devices are outlined in Table 3.

**Table 3.**
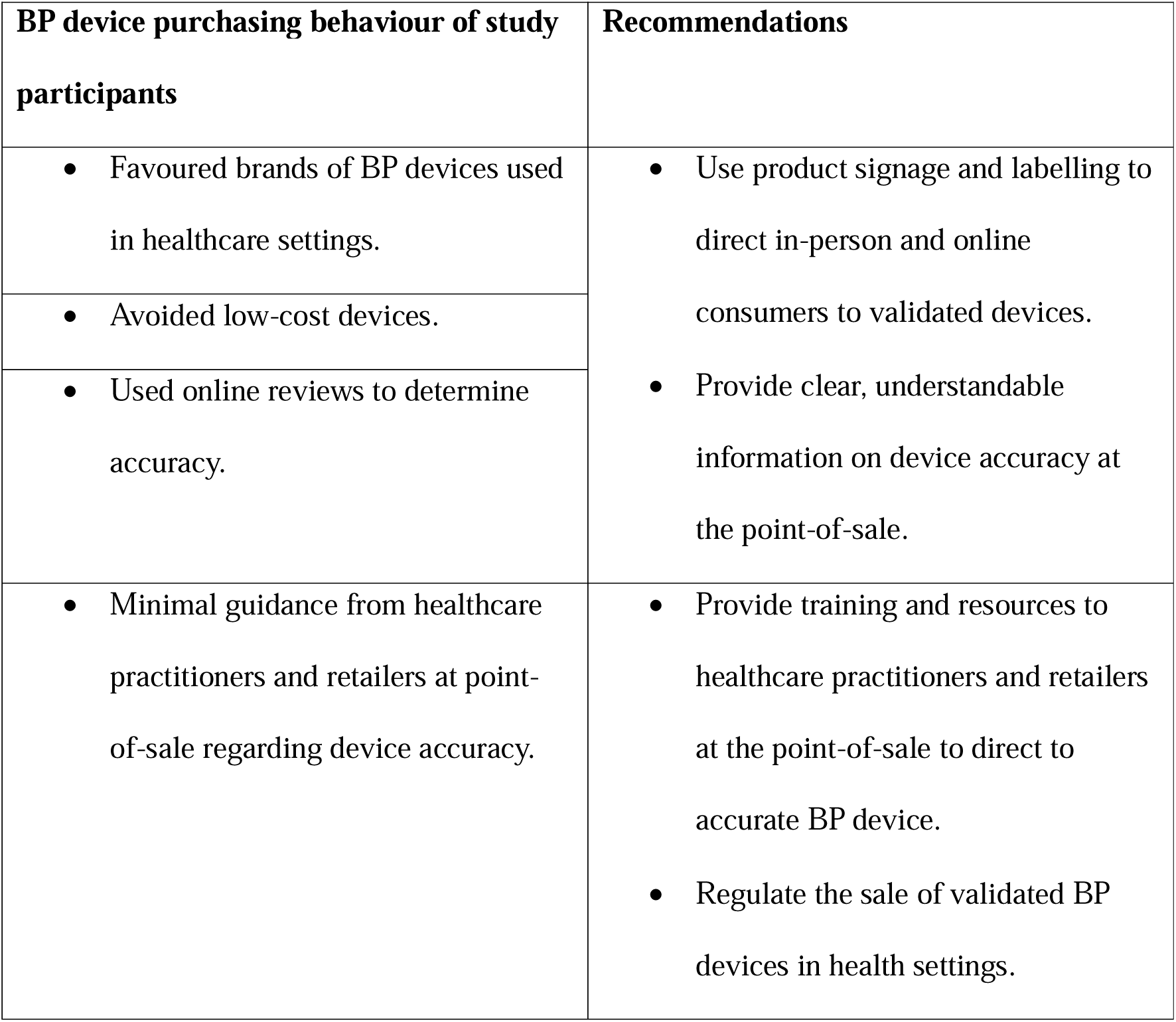
Recommendations to direct consumers to obtain a validated blood pressure device for use at home.

Using a validated BP device is recommended to reliably obtain accurate BP readings to inform BP clinical decision making (9–11) for BP management and control (2, 33–35). However, most devices available to purchase are not validated (9, 16, 36), including in Australia where only a small proportion of devices available to purchase from pharmacies and online are validated (24% & 6%, respectively) (16). Despite this, our study found that around half of the devices used by Australians for HBPM were validated, which is higher than the availability of validated BP devices previously observed in the Australian market. While previous studies have identified demographic characteristics associated with BP device ownership (37), this study builds on this evidence by identifying the factors influencing device selection. Participants in our study employed strategies at point-of-sale to select BP devices which they believed to be accurate, including using cost to benchmark the accuracy of a device, selecting a recognisable brand used in healthcare settings and buying from pharmacy as a trusted health setting. Additionally, while participants reported that they valued the advice of healthcare practitioners regarding BP device selection, advice about validation status was not received when obtaining a device. This suggests that adults can be directed to select a validated BP device by providing consumers with guidance in healthcare settings such as at physician clinics when HBPM is recommended by GPs and in pharmacies at the point-of-sale of devices.

Strategies at the point-of-sale to improve the availability and use of validated BP devices have been implemented in other settings internationally. Hypertension Canada have promoted greater uptake of validated BP devices, particularly in pharmacy, through the application of stickers of endorsement on the packaging of validated devices and through an online ‘Hypertension Canada Listing’ of validated devices (38). Research data indicates that 90% of devices stocked in Canadian pharmacies are validated (39), suggesting that increasing the visibility of validated BP devices may impact the behaviour of pharmacy owners and support consumers to obtain a validated BP device for use at home. In England, previous research among participants recruited from a GP hypertension registry identified that 50 different device models were used, of which 69% were validated despite only 22% of devices that are available for purchase online in this region being validated (19, 36). Similar results were found by our study, where 86 device models were used (51% validated). Strategies used in England which may contribute to higher uptake of validated BP devices despite low market availability include organisations such as the British Heart Foundation outlining the importance of BP device validation, listing the validated BP devices recommended for home use and exclusively selling validated BP devices on their online store (40, 41). These international examples illustrate how strategies which enable the easy identification of validated BP devices and encourage pharmacies to stock validated devices may improve the uptake of validated BP devices for home use among consumers. The findings of this study, which show that 69% of devices were purchased at a pharmacy with little guidance from health professionals, indicate that such strategies would be effective to direct Australian adults to use validated BP devices. A cost benefit analysis of achieving universal use of validated devices estimated a return of $2.7-$3.2 AUD through improved hypertension diagnosis and management after accounting for expenditure for healthcare practitioner training, policy change and industry engagement activities (42), supporting the potential of such strategies.

Most participants of this study purchased their BP device from a pharmacy, identifying this as a key setting to support the use of validated BP devices. Pharmacies are a highly accessible and frequently visited health setting for long-term BP management, particularly for adults using anti-hypertensive medication (43). Pharmacists play an important role in a team-based care approach to BP management, which achieves improved BP outcomes at a reduced cost through maximising healthcare practitioner scope of practice and the delivery of patient education outside of the GP office (44, 45). However, a survey of Australian pharmacists found that more than half believed that all BP devices sold in pharmacies had passed validation testing and most were unable to identify a validated BP device (46). This was supported by the findings of our study, as participants described receiving guidance from pharmacists regarding retail availability, technological features and ease-of-use of BP devices, rather than information about device validation as an indication of accuracy. These findings indicate that although pharmacies are an important healthcare setting for BP management, pharmacists do not have the knowledge to advise adults on key BP management topics. Pharmacists require capacity building through training and resource provision to enable them to support adults in self-monitoring of BP and fulfil their important role in a team-based care approach to BP management.

Achieving widespread changes to the availability and subsequent use of validated BP devices may require changes in regulation that mandate the exclusive sale of validated BP devices in health settings. The present study found that adults and healthcare practitioners did not attempt to identify or recommend validated BP devices for home use and instead employed the strategies to select a BP device from the range of devices available. In particular, participants felt that pharmacies were a trustworthy source to purchase a BP device from and expressed trust in the device brands used in healthcare settings. This highlights the responsibility of healthcare professionals to provide consumers with accurate health products and equipment. As accurate BP measurement is integral to BP control, regulatory bodies should consider the implications of selling non-validated BP devices on the cardiovascular health of customers and impose stricter regulation to ensure the sale of validated BP devices in health settings. Increasing the availability of validated BP devices in health settings such as pharmacies is a powerful tool to encourage uptake of these devices, particularly in the absence of consumer strategies to identify validated devices.

### Strengths and limitations

This study is strengthened by its mixed methods design with complementary interview data to provide a deeper understanding of the factors that influence the BP device used at home for BP measurement. However, the study was conducted in only one country (Australia) and is limited by the relatively small sample size and low ethnic diversity of the participant sample, as 90% were White. Adults from culturally and linguistically diverse communities may consider unique factors when selecting a BP device. In addition, adults with a greater interest and perceived competence in HBPM may have been more likely to participate in the study. These factors, as well as other cultural barriers to healthcare access, should be included when considering how to support adults to obtain validated BP devices. The cost of BP devices reported by survey participants was extracted using a methodology reflecting the cost-saving purchasing strategy described in participant interviews and may not have been the exact cost incurred by the participant. However, the cost findings are in line with previous research from Australia (16). This study has not explored experiences surrounding the selection of an appropriately sized cuff when obtaining a BP device. This is another important factor that impacts BP measurement accuracy and should be explored by future studies (47).

## Conclusions

This study found that only half of BP devices used by participants for HBPM in Australia were validated. Despite accuracy being stated as the most important factor when selecting a BP device, participants did not receive sufficient support from healthcare providers to clearly identify a reliable, accurate device. Instead, participants used device brand, cost and online reviews to select an accurate BP device. Findings from this study highlight the need to support consumers to select validated BP devices by training pharmacy staff to direct consumers to validated BP devices and by providing clear labelling at point-of-sale. BP measurement using a validated device is an important part of ensuring accurate HBPM to support clinical decision making for BP management. This study draws from the real experiences of study participants to recommended strategies to improve uptake of validated BP devices for HBPM.

## Data Availability

Data from this study are available upon reasonable request to the corresponding author.

## ACKNOWLEDGEMENTS

We sincerely thank Carol Batt and John Stevens for their contributions to this work as consumer advisors.

## SOURCES OF FUNDING

Niamh Chapman is supported by a National Heart Foundation of Australia Postdoctoral Research Fellowship [2023-2024, #106657]. DSP is supported by a National Health and Medical Research Council Investigator Grant (GNT2018077) and is an Honorary Future Leader Fellow of the Heart Foundation of Australia (106618). Aletta E Schutte is supported by an NHMRC Investigator Leadership Grant [2023-2027, APP2017504).

## CONFLICTS OF INTEREST

Aletta E Schutte has received speaker fees from Omron, Medtronic, Aktiia, Servier, Sanofi, Novartis and is advisory board member for Skylabs and Abbott.

Ross T Tsuyuki has received investigator-initiated, arm’s length research grants from Merck, Sanofi, AstraZeneca, and Pfizer. He has been a consultant for Merck, Shoppers Drug Mart, PharmaSmart, and Emergent Biosolutions.

NC, DSP, EC have no COI to declare.

## NON-STANDARD ABBREVIATIONS AND ACRONYMS

BP: blood pressure
GP: general practitioner
HBPM: home blood pressure measurement HSC: high school certificate

## SUPPLEMENTARY MATERIAL

Table S1-S8

## Notes

### Author Declarations

This study was approved by the Human Research Ethics Committee of the University of Tasmania (H0028867) and informed consent was provided by all participants prior to participation.

